# Estimating Realized Access to Obstetric Care in Georgia: A Discrete Choice Modeling Analysis

**DOI:** 10.1101/2025.11.07.25339779

**Authors:** Jingyu Li, Margaret Carrel, Stephanie Radke, Lauren N. Steimle

**Author notes:** **Corresponding Author** Lauren N. Steimle, PhD, Address: 755 Ferst Dr NW, Atlanta, Georgia 30318.

## Abstract

The ongoing closure of obstetric units in the United States poses challenges to maternal care access, especially in rural communities. These closures leave patients with limited options for obstetric services and may force them to travel longer distances or rely on low-volume facilities, both associated with adverse maternal outcomes. While policymakers have proposed interventions to support obstetric care in underserved areas, evaluating their potential impact requires understanding how birthing patients select among available facilities and how those selections affect delivery volume and access patterns. This study develops a revealed preference discrete choice modeling framework to estimate realized access to obstetric facilities—the extent to which patients actually use available obstetric services—based on patient characteristics and facility attributes. Using 2016-2019 birth records in the state of Georgia, we developed three discrete choice models: a distance-only model, a multinomial logit model based on facility attributes, and a latent class model that considers heterogeneous weights patients place on facility attributes. We evaluated model performance in predicting low-volume facilities and regional access. The latent class model outperformed others and identified two patient classes: “less distance-sensitive” patients (34.5%) and “more distance-sensitive” patients (65.5%) with differing sensitivity to distance and levels of care. Our findings reveal regional patterns where patients disproportionately seek care far away or at low-volume facilities, and how these patterns may change as obstetric units close. We highlight the practical use of our model for forecasting underutilized facilities and informing policies to better distribute patient volume across the region.

**Highlights:**

- We analyzed how pregnant people in Georgia, USA sought obstetric care using statewide birth records from 2016-2019
- We developed and compared three discrete choice models (distance-only model that assumes patients select nearest obstetric facilities, multinomial logit model, and latent class model) that considered patient characteristics and facility attributes to predict patients’ utilization of birth facilities and validated these models using out-of-sample datapoints.
- Travel distance, facility level of care, rurality of facility location, and health system affiliation significantly influenced patients’ selection of delivery facilities. We also identified two classes of patients who varied in their social determinants of health and placed heterogenous weights on facility attributes that affected their selection of obstetric facilities.
- Targeted interventions to improve maternal care access in underserved areas should consider patients’ underlying social determinants of health, as geographic proximity alone is not predictive of patients’ selection of delivery facility.
- We demonstrate how our model can be applied to forecast low-volume obstetric facilities and access to care and to inform policies that avoid underutilized facilities and better distribute patient volume across the region.

## 1. Introduction

Closures of obstetric (OB) units in the United States (U.S.) present a critical challenge to maternal care access, particularly in rural communities. Between 2006 and 2020, over 400 OB units shut down nationwide [1]. The disappearance of OB units has been particularly devastating in rural areas, where 267 facilities, nearly a quarter of all rural OB units in the U.S., stopped providing childbirth services between 2011 and 2021 [2]. These closures have left rural populations with increasingly limited options for OB care. As a result, some patients, especially those residing in rural areas, must travel long distances to reach a delivery facility. Others may rely on a more local option for care, which is often a low-volume facility for rural populations. Far travel distance and delivery in low-volume rural facilities are both associated with increased rates of maternal morbidity and mortality [3, 4].

The reasons for OB unit closures have been well-documented. Hospital administrators commonly cite financial viability and workforce shortages as reasons for closing OB units [5]. Operating an OB unit incurs large fixed costs each year, which hospitals seek to make up through reimbursements. However, facilities with low patient volume may struggle to offset the fixed cost associated with the OB service lines [6], and thus to maintain sufficient patient volume for financial viability and patient safety. This challenge is compounded by patient *bypass*, as estimates suggest between 11% and 55% of patients choose to bypass their local birthing facility in favor of one farther away [7, 8]. These decisions may reflect a preference for higher-quality or more risk-appropriate care [9, 10], dissatisfaction with local services [11], or desire for culturally concordant care [12, 13]. When closures occur, they can trigger a negative cycle as departing obstetricians and nurse-midwives further exacerbate the existing workforce shortages.

Thus, as policymakers seek to address gaps in access to OB care and to support rural OB facilities, they must understand how birthing people make decisions about where to deliver, given the birthing facilities available. Traditional measures of access to OB care, such as geographic proximity to birthing facilities, overlook behavioral dimensions of access—how patients respond to available options, as patients may bypass nearby facilities in response to perceived quality, distance, or facility attributes. Simply assessing historical volume trends is insufficient for guiding forward-looking initiatives, as this approach fails to capture how patients will make selections of their birthing facility as the available OB units change. To understand how patients seek birthing facilities and how these choices may change in response to closures or consolidations, we use revealed preference (RP) discrete choice modeling to understand *realized access* to OB care in the state of Georgia, that is, where patients actually seek care as opposed to simply how far away they live from the closest birthing facility.

### 1.1. Literature Review

#### 1.1.1 Access to OB care

Access to care is a multidimensional concept. In this study, we focus specifically on geographic access, which is often assessed through two distinct perspectives: *potential access* and *realized access* [14]. Potential access refers to the spatial availability of care services, while realized access captures the actual utilization of those services by patients given their location relative to care. In the context of OB care, potential access is typically measured by indicators such as provider-to-population ratios, travel distance to the nearest delivery facility, or the presence of a birthing facility in some geographic unit. For example, the “maternity care desert” framework developed by the March of Dimes identifies counties with limited or no access to OB services based on the availability of maternity care providers and facility-based OB services (March of Dimes). Another measure is the distance to facilities offering critical care OB (CCO) services [15, 16]. While these measures are valuable for identifying structural gaps, particularly in underserved areas, they fail to capture how individuals use available services. Such measures assume patients will seek care at the nearest facility and overlook the variations in preferences, perceived quality, transportation access, and other social determinants of health.

In contrast, realized access reflects patients’ actual use of healthcare services when seeking care. For instance, some patients bypass their closest OB facility to seek care at a farther OB unit. Recent studies have documented this behavior empirically in the U.S. Thorsen et al. reported that 11% of births in Montana occurred at facilities farther from patients’ closest labor and delivery (L&D) units [8]. In Iowa, more than half of patients bypassed the nearest OB provider when delivering [7]. Several factors are associated with increased likelihood of bypassing including complex medical diagnoses, higher income, younger age, private insurance coverage, and expanded Medicaid eligibility [7, 9, 17–21]. Conversely, patients with lower income, inadequate prenatal care, or public insurance are more likely to deliver locally [22, 23].

Despite these insights, existing studies often fall short in capturing the full complexity of patient decision-making. Many rely on descriptive or regression-based analyses that do not adequately model the choice process or account for variation across individuals. In addition, many of these studies do not incorporate facility-level and patient-level characteristics simultaneously, or fail to validate findings on out-of-sample data. Their utility for informing policy design and intervention planning needs further investigation.

#### 1.1.2 Background on discrete choice modeling

Understanding what drives patients’ selection of birthing facilities is a key component for accurately estimating patients’ realized access to OB care and thus identifying regions that may warrant targeted support. In recent years, discrete choice modeling has emerged as an approach to analyze such decisions. Discrete choice models, grounded in behavioral theory, estimate the probability that a decision-maker (DMs) selects a particular option from a defined set of distinctive alternatives and are well-suited for forecasting choices under hypothetical scenarios [24]. They have been applied across a range of contexts, including transportation and travel behavior [25, 26], marketing and retail [27, 28], public policy [29–31], and operations management [32]. A detailed review of this subject is provided by Train 2009 [33].

A central distinction in discrete choice modeling is between revealed preferences (RP) and stated preferences (SP). RP are derived from actual choices DMs have made in real-world settings. In contrast, SP are elicited through hypothetical scenarios such as surveys or experiments where individuals are asked to indicate their choices [34]. RP data reflect real behavior but may lack variation in key variables as those data are pre-collected, while SP data can be designed to capture such variation but the consistency of choices may be influenced by how a question is asked.

The theoretical foundation of discrete choice modeling lies in utility maximization theory, which posits that individuals act rationally to maximize their utility, subject to constraints. In the context of discrete choices, utility is composed of an observable component (based on measurable attributes of the alternatives and DM characteristics) and an unobservable random component. Multinomial logit (MNL) model is one of the most widely used discrete choice models and the basis for many other random utility maximization (RUM) models. The key assumption of MNL model is that the random component is independently and identically distributed with a Gumbel distribution. This assumption allows for a closed-form choice probability but imposes the independence of irrelevant alternatives (IIA) property, which may be restrictive especially when alternatives share unobserved similarities [35]. To address the limitations of MNL models, latent class (LC) model is introduced to handle unobserved heterogeneity in individual taste. This model is built on the idea of a finite set of distinct “patient classes,” allowing attribute level coefficients to remain consistent within each class but vary between classes [36].

Prior literature has demonstrated discrete choice modeling as a promising approach to understand patients’ choices in health services research. Studies have consistently identified travel distance and quality of care as key factors affecting patients’ healthcare choices in both U.S. and international contexts [37–41]. More recent studies have emphasized that patients are not a homogeneous group, and that preferences vary by demographic, socioeconomic, and clinical characteristics [37, 42–45]. For example, several studies have identified subgroups of patients who are more willing to travel longer distances to access higher-quality care [37, 45].

To our knowledge, the most closely related studies to ours are that of Hwang et al., 2017 and Jang et al, 2017 who used MNL models to estimate patients’ realized accessibility to OB care in South Korea [46, 47]. Others have formulated a gravity-based nonlinear optimization model to estimate access at rural OB hospitals in Montana [48]. However, these studies typically rely only on facility-level attributes and do not incorporate patient-level heterogeneity, which limits their ability to capture variation in behavior that may be driven by patient-level characteristics. Another study used a latent-class choice model to understand the impact of social determinants of health on the choice of delivery facilities among pregnant women in low-income country, which is a very different context for OB care and provides limited understanding into facility closures in the U.S. [44].

Our study presents a novel way to understand patients’ delivery facilities choices that includes both facility and patient-level covariates in the unique context of OB unit closure. This RP approach allows us to account for heterogeneity in how patients evaluate facility attributes. We further validate our model using out-of-sample data and assess performance on outcomes directly relevant to access measure—such as proximity to the delivery facility and use of low-volume facilities.

### 1.2 Contributions

In this paper, we present a novel application of discrete choice modeling to estimate and predict realized access to birthing facilities. To do so, we conducted a revealed-preference case study in the state of Georgia. We focus on a single-state study because although general healthcare regulations are imposed at the national level under the U.S. healthcare governance structure, there is substantial variation across states, as many specific decisions are made at the state level. Georgia was selected as a highly relevant case study due to its high rates of maternal morbidity and mortality and large rural population. Our study makes several contributions:

- First, we demonstrate that realized access differs significantly from access defined by geographic proximity alone. By incorporating factors such as facility level of care, health system affiliation, and location into choice modeling, we provide a more accurate representation of how patients select delivery facilities in the U.S. context. This result further suggests the limitations of proximity-based forward-planning approaches and emphasizes the importance of considering patients’ actual choices when evaluating maternal care access.
- Second, we assess model performance against several critical measures, including (1) geographic proximity between the patients’ residence and delivery facility, and (2) whether patients deliver at low-volume hospitals, which are associated with poorer maternal outcomes. These validation steps demonstrate the practical utility of our model in identifying geographic areas and subpopulations where targeted intervention to improve OB access are most needed.
- Third, we distinguish different types of patients that weigh facility attributes differently. This finding suggests interventions to improve maternal care access should be tailored to the needs of different patient subgroups.
- Lastly, we make innovative use of statewide birth records as an RP dataset. These records provide a high-resolution, population-level view of patients’ actual facility selection of birthing facilities. Such practice can be easily adapted for use in other states with different healthcare regulatory environments or patterns of healthcare access disparities.

## 2. Methods

### 2.1 Sample

We analyzed birth records for all births to Georgia residents from January 1^st^, 2016, to December 31^st^, 2019, in the state of Georgia. Figure 1 provides an overview of our exclusion criteria of birth records. First, we excluded deliveries that had missing records of residence or birth facilities. Then, we excluded deliveries that involved a maternal transport as these cases do not reflect patients’ original selection of delivery facilities. Additionally, we excluded deliveries at non-OB facilities that are not included in the 2017 Georgia guidelines for perinatal regionalization, since we expected that these cases likely reflect situations where delivery was urgent and there was limited to no patient control over birth facility selection [49]. Lastly, we further excluded deliveries that occurred more than 120 miles away from the birthing person’s residential address, as selecting a facility at such a distance is highly unlikely under typical circumstances. We suspected that such cases were likely due to emergencies that occurred while the patient was traveling or to high-risk, complex deliveries requiring specialized care available only at distant facilities. Since 99.75% of all deliveries occurred within 120 miles of the birthing person’s residence, excluding these observations is expected to have minimal impact on the analysis. The final sample included 489,512 deliveries. The Georgia Tech Institutional Review Board approved this study (Protocol H23091). We fitted our models using 2016-2018 birth records in Georgia and validated our models using 2019 birth records.

**Figure 1.**
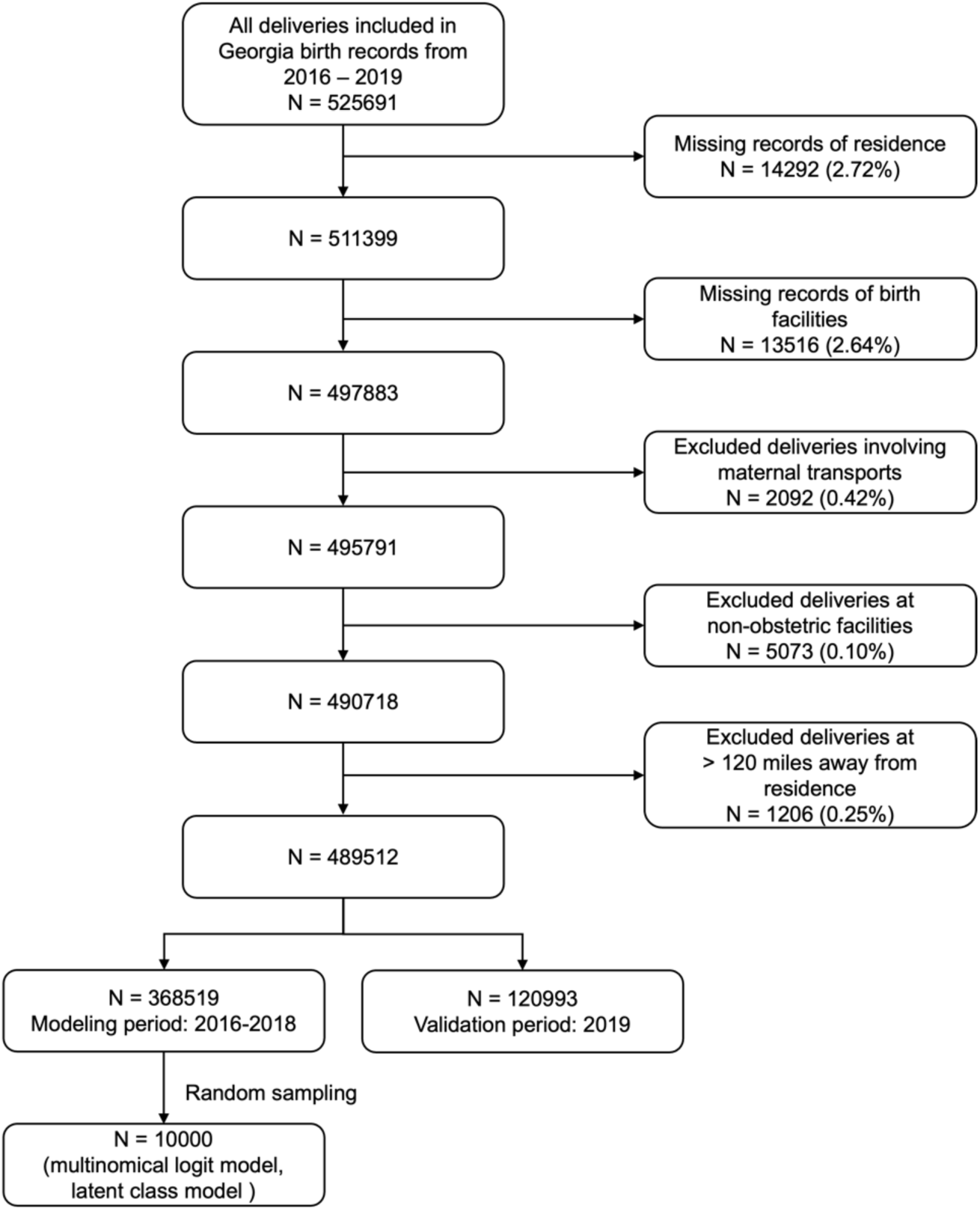
Data inclusion criteria.

### 2.2 Discrete choice modeling framework

In this section, we describe the data used to fit our discrete choice models and our modeling process.

#### 2.2.1. Factors included in discrete choice models

We considered both facility-level attributes and patient characteristics as factors in our discrete choice models. For patient characteristics, we included all medical risk factors (for example, number of fetuses, presence of gestational diabetes, and or hypertension) and sociodemographic variables (for example, age, race, and ethnicity) available in the birth records. A detailed description of those variables can be found in **Appendix Table A1**. To guide selection of facility-level attributes, we adapted a conceptual framework that identified seven domains of provider characteristics that influence patient decision-making [46, 50]. Specifically, we considered: availability (availability of the provider, such as open hours and variety of options which might be constrained by patients’ insurance plan), accessibility (whether the provider is easily accessible by patients’ own or public transportation), type and size, staff, organization of care, cost, and provider demographics (demographics of individual doctors). Due to data availability, we excluded cost and provider demographics from model fitting.

For availability, we assumed patients were free to choose from the set of all operating OB facilities within 120 miles of their residence. As proxies for accessibility, we used road distance between patients’ residences and birthing facilities, as well as whether the facility is in an urban area. We calculated road distance between patients’ residence, represented by the 2020 census block group population-weighted centroids and OB facilities where deliveries occurred using Open Source Routing Machine (OSRM) API in Python. As a proxy for type and size, we used *level of care* which represents a facility’s capabilities and resources for handling maternal and fetal risks [51]. To represent staff, we included whether a facility offered midwifery services, defined as having at least one delivery attended by a midwife. For the organization of care, we included two additional factors from public sources, including Baby-Friendly designation [52], which indicates that a facility adheres to evidence-based practices for breastfeeding and maternal-infant care, and health system affiliation of each facility, which was obtained for each facility each year.

To avoid multi-collinearity among factors, we calculated variance inflation factors for all candidate variables and confirmed that none of them were correlated with each other (**Appendix Table A2**).

#### 2.2.2 Multinomial Logit Model

The MNL model represents a patient *i*’s true utility of OB facility *j* (*U_ij_*) among a set of available OB facilities (*J*) as the sum of two parts: an observable utility (*V_ij_*) and a random error term (*∈_ij_*). The observable utility can depend on both facility-related attributes (*X_ij_*) and patient-related characteristics (*X_i_*), while the error term follows a Gumbel distribution [35]. Let *D*(*i*) denote patient *i*’s decision, then the probability of patient *i* choosing facility *j* (*P_ij_*) can be calculated using a closed-form expression.

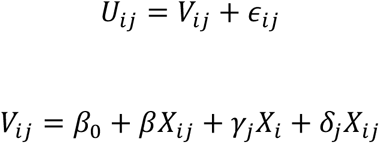

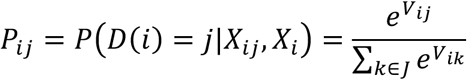

Since choice sets available to each patient can vary, we do not estimate any alternative-specific coefficients (*β*_o_ = *γ*_j_ = *δ*_j_ = 0 ∀ *j* ∈ *J*).

To examine whether or not a LC model might be appropriate due to heterogeneity in patient preference weights, we first fitted MNL models for subgroups of patients. Specifically, we estimated separate models for groups defined by key patient characteristics, such as race, ethnicity, education level, payer status, and rural versus urban residence. For instance, we fitted separate models for rural and urban patients and determined if there were differences in their coefficients. By comparing results for rural and urban patients, we could evaluate whether facility attributes such as proximity to residence and level of care influenced facility choice differently depending on where patients lived.

#### 2.2.3 Latent Class Model

The LC model takes into account heterogeneity in the weights patients might place on the factors that influence their selection. The model includes two components: a class membership model and a class-specific choice model. Let *Q*(*i*) denote the index of the class of the individual *i*. The class membership model calculates the likelihood of a patient in each class *c* based on patient characteristics *P*(*Q*(*i*) = *c*|*X*_*i*_). Meanwhile, the class-specific choice model estimates the likelihood of selecting an OB facility given each class *c*. Following the same notation, the probability of patient *i* choosing facility *j* can be calculated as followed.

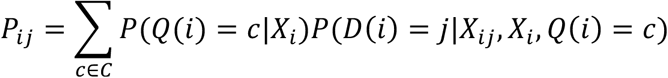

For this analysis, we began by including all relevant sociodemographic and medical risk factors in our class membership model. A detailed description of the tested variables is included in **Appendix Table A1**. To refine our model, we performed variable selection to identify variables that were statistically significant and distinct across latent classes. We also tested multiple number of latent classes and iteratively examined each for model convergence, statistical significance, model fit, and interpretability. Since log likelihood always improves as the number of latent classes increases, we used Akaike’s Information Criteria (AIC) and Bayesian Information Criterion (BIC), which have been suggested in prior literature, to compare model fit during model selection process [53, 54].

Due to memory limitations of the working protected health data environment in which the birth records were stored, we randomly sampled 10,000 datapoints from 2016-2018 records to fit MNL and LC models. To ensure the robustness of our sampling, we also estimated a simplified model using all available data from 2016–2018. This model included only three predictors: road distance, facility level of care, and whether the facility is in an urban area. The coefficients estimated from the sampled dataset, which have slightly wider 95% confidence intervals (CIs) due to the smaller sample size, still narrowly encompass the corresponding estimates from the full dataset (**Appendix Tables A3 & A4**). This suggests that our downsampling approach did not significantly affect the estimated relationships, because the effects of the predictors observed in the sampled data are statistically compatible with those in the full dataset.

### 2.3 Out-of-sample Validation

We compared and validated our fitted discrete choice models using 2019 data against the distance-only model which assumes that patients choose the closest facility. We used four evaluation metrics to compare these models. First, we used logarithmic score, which is a widely adopted strictly proper scoring rule, to assess how well each model predicted individual selection of OB facilities [55, 56]. This metric measures the choice prediction performance by comparing the predicted choice probabilities with the actual observed choices. Logarithmic score is unbounded from below, and a higher score value indicates better probabilistic prediction. Second, we compared the performance of predicting low-volume facilities of each model, measured by accuracy, specificity, sensitivity, and F1 score. We defined a low-volume facility as an OB facility with less than 500 annual delivery volume [57]. We also conducted a sensitivity analysis by applying alternative thresholds of 250, 300, and 400 annual delivery volume per year.

In addition to evaluating predictive accuracy, we used the models to forecast key dimensions of geographic access to OB care. However, access to OB care can be conceptualized and quantified in multiple ways, and there is currently no standardized measure or metric exists in the U.S. Distance to facility has been used in previous literature and is linked to adverse maternal outcomes [3, 58, 59]. Therefore, we compared the predicted average travel distance to the chosen facility at the census tract level. Regions with higher predicted distances may indicate areas with reduced access. Another important dimension of access to care is the facility volume due to volume outcome relationships [4]. We then compared the predicted percentage of patients delivering at low-volume facilities (defined as predicted to have less than 500 annual deliveries) with the observed percentage (in facilities that actually had less than 500 annual deliveries). Finally, we calculated the Spearman’s correlation coefficient between the predicted and observed percentages across census tracts to assess model performance in capturing spatial variation in access.

To facilitate interpretation across models, all facility attributes were translated into “road distance equivalent” (RDE) units by dividing each attribute’s coefficient by the (negative) coefficient of the road distance. This allows the impact of non-distance attributes to be interpreted as equivalent reductions in travel distance. RDE simplifies comparisons across models and supports assessing the relative importance of each facility attribute in terms of travel burden [60].

All discrete choice models were fitted in R (version 4.4.1) using package Apollo (version 0.3.3) [61]. All other analyses were performed in Python (version 3.11.9). We used a p-value of 0.05 as the significance threshold throughout all analyses.

## 3. Results

### 3.1 Sample

A total of 489,512 deliveries met the inclusion criteria shown in Figure 1. Of these, 368,519 deliveries occurred between 2016 and 2018 (during modeling period) and 120,993 occurred in 2019 (during validation period). The analysis included 79 OB facilities, two of which closed their OB units during the study period—one in 2017 and the other in 2018. Among 77 operational OB facilities in 2019, 19 (24.68%) had annual volume less than 500, 18 (23.38%) had annual volume less than 400, 12 (15.56%) had annual volume less than 300, and 9 (11.69%) had annual volume less than 250. Descriptive statistics for the included facilities are presented in **Table 1**.

**Table 1.** Descriptive statistics of facilities.

|  | Total<br>(N = 79) |
| --- | --- |
| **Average Annual Volume** |  |
| Mean (SD) | 1550 (2020) |
| Median [Min, Max] | 1010 [109, 15800] |
| **Level of Care, N (%)** |  |
| Birth Center | 2 (2.5) |
| 1 | 26 (32.9) |
| 2 | 26 (32.9) |
| 3 | 18 (22.8) |
| RPC | 7 (8.9) |
| **RUCC Classification, N (%)** |  |
| Rural | 9 (11.4) |
| Suburban | 19 (24.1) |
| Urban | 51 (64.6) |

To ensure the modeling sample was representative of the population, we compared the distribution of patient characteristics and facility volumes in the sampled dataset and the full dataset using Chi-square test of independence. We observed no significant difference, suggesting that our sampled dataset had good fit of the full dataset.

### 3.2 MNL model

**Table 2** presents the results of the estimated MNL model. RDE interpretation for each variable is detailed in **Table 4**. All facility-related attributes were statistically significant and were retained in the final model. The negative coefficient for road distance indicates that patients viewed longer travel distances unfavorably. Patients preferred facilities offering higher levels of care compared to level I facilities, with an increase in utility observed from birth centers to level III facilities (p < 0.001). When interpreted in RDE, patients would be equally willing to choose a birth center that is 20.39 miles closer, an RPC that is 9.09 miles farther, a Level II facility that is 6.79 miles farther, a Level III facility that is 13.21 miles farther, than a Level I facility. In addition, patients were willing to travel an additional 5.99 miles for facilities located in urban areas, 1.54 miles for those offering midwifery services, 2.14 miles for Baby-Friendly designated hospitals, and 3.76 miles for facilities affiliated with a multi-facility health system.

**Table 2.**
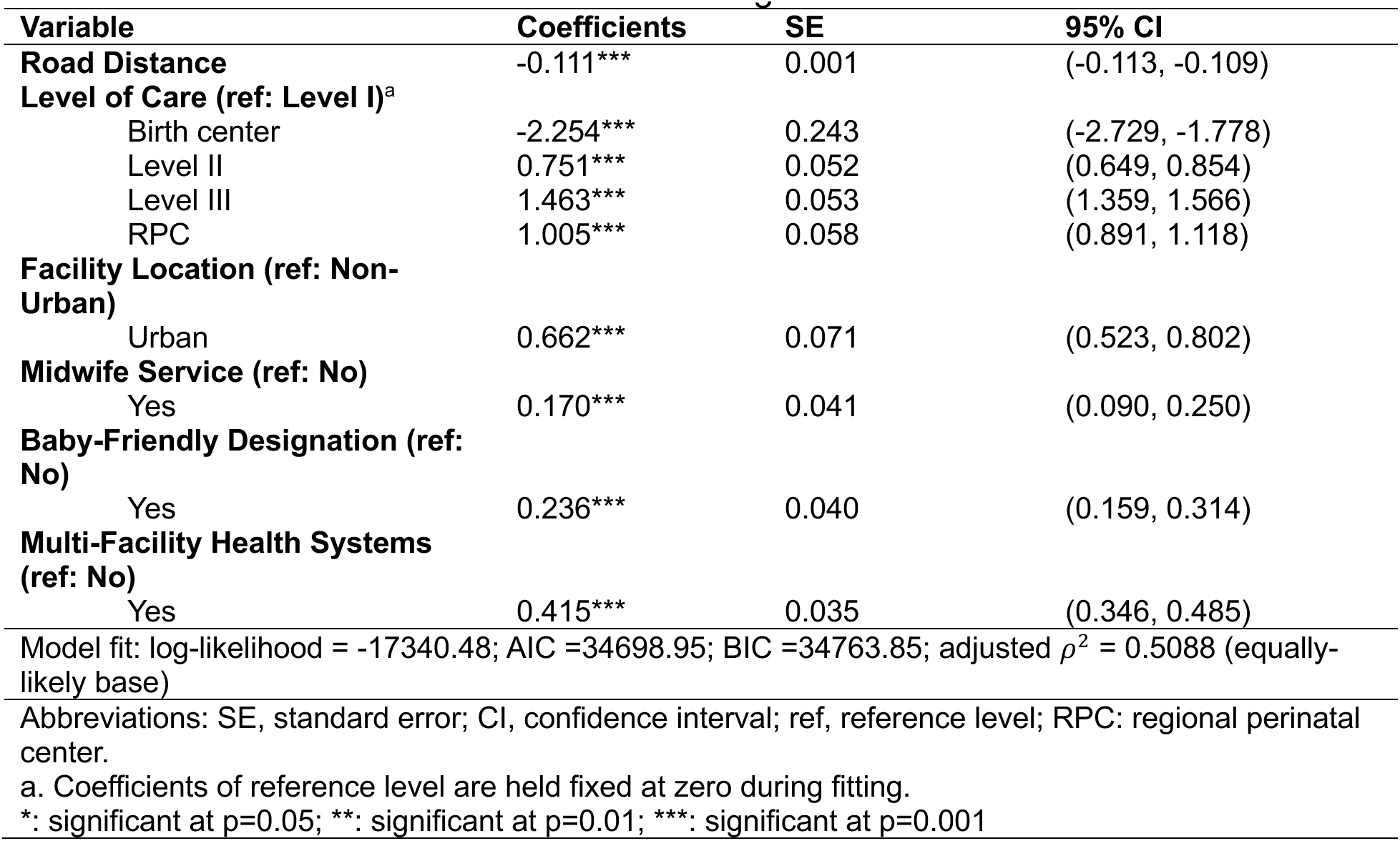
Estimated results for the multinomial logit model.

| Variable | Coefficients | SE | 95% CI |
| --- | --- | --- | --- |
| **Road Distance** | -0.111*** | 0.001 | (-0.113, -0.109) |
| **Level of Care (ref: Level I)<sup>a</sup>** |  |  |  |
| Birth center | -2.254*** | 0.243 | (-2.729, -1.778) |
| Level II | 0.751*** | 0.052 | (0.649, 0.854) |
| Level III | 1.463*** | 0.053 | (1.359, 1.566) |
| RPC | 1.005*** | 0.058 | (0.891, 1.118) |
| **Facility Location (ref: Non-Urban)** |  |  |  |
| Urban | 0.662*** | 0.071 | (0.523, 0.802) |
| **Midwife Service (ref: No)** |  |  |  |
| Yes | 0.170*** | 0.041 | (0.090, 0.250) |
| **Baby-Friendly Designation (ref: No)** |  |  |  |
| Yes | 0.236*** | 0.040 | (0.159, 0.314) |
| **Multi-Facility Health Systems (ref: No)** |  |  |  |
| Yes | 0.415*** | 0.035 | (0.346, 0.485) |
| Model fit: log-likelihood = -17340.48; AIC =34698.95; BIC =34763.85; adjusted $\rho^2$ = 0.5088 (equally-likely base) | | | |
| Abbreviations: SE, standard error; CI, confidence interval; ref, reference level; RPC: regional perinatal center. |  |  |  |
| a. Coefficients of reference level are held fixed at zero during fitting. |  |  |  |
| *: significant at p=0.05; **: significant at p=0.01; ***: significant at p=0.001 |  |  |  |

Fitted MNL models by subgroups are shown in **Appendix Figure B1-B5**. The results further confirmed that patients placed heterogeneous importance weights on facility attributes. For example, patients with commercial insurance and Medicaid would be equally willing to choose a level III facility located 17.78 and 10.15 miles further than a level 1 facility, respectively. Therefore, an LC model, which can account for exogeneity in patients might better address that there is a dimension to the input data that is not captured.

### 3.3 LC model

#### 3.3.1 Estimates

We started with 2-class LC models and included all potential patient-level variables as detailed in **Appendix Table A1**. We noticed that the coefficient for birth center approached negative infinity since there were only 2 birth centers and very few deliveries occurred at such facilities. To resolve this convergence issue, we assigned the MNL coefficient for birth centers and held it as fixed during model estimation. We did not test LC models with 4 classes or more because the estimation for 3-class models was unstable and poor (**Appendix Table B1**).

**Table 3** shows the results of the final estimated LC model, which had 2 classes. Their RDE interpretation is detailed in **Table 4**. Coefficients and their 95% CIs for detected latent classes are visualized in **Figure 2**. We accepted this model because it offered good interpretability and had the best model fit (AIC = 33686.81; BIC = 33867.07; adjusted *ρ*\* = 0.5231) among all candidate models that identified statistically significant and distinct patient-level variables across latent classes. We observed notable differences in class-specific choice models across two identified latent classes. These differences confirmed that distinguishing heterogeneity in importance weights for each class was important. Specifically, road distance to OB facilities had more negative coefficients in class 1 compared to class 2, which shows that patients in class 1 viewed longer distances to facilities less favorably. To distinguish between these classes, we refer to class 1 as “more distance-sensitive,” and class 2 as “less distance-sensitive”. Both groups favored facilities with higher levels of care compared to level I facilities, as suggested by positive coefficients for Level II, Level III, and RPCs for both groups. When interpreted in terms of RDE, Class 1 patients would be equally willing to choose a birth center that is 12.89 miles closer, a Level II facility 5.27 miles farther, a Level III facility 6.74 miles farther, and an RPC 3.17 miles farther than a Level I facility. In contrast, Class 2 patients would accept significantly longer travel distances: 33.74 miles closer for a birth center, 17.43 miles farther for a Level II facility, 45.43 miles farther for a Level III facility, and 47.24 miles farther for an RPC. This suggests that “less distance-sensitive” patients showed much stronger preference to level III facilities and RPCs compared to “more distance-sensitive” patients. Relying solely on a single-class model as in MNL might have underestimate these effects, as the computed sample-wide “average” coefficient could underestimate preference for level III and RPCs for “less distance-sensitive” patients.

**Figure 2.**
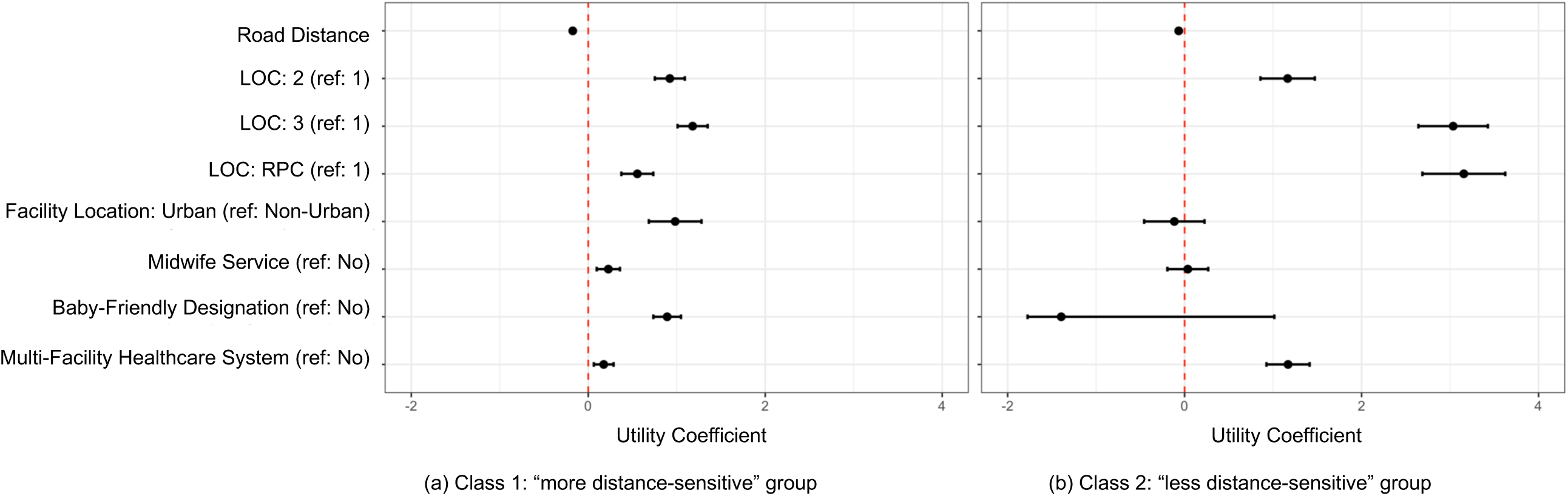
Estimated results of the LC model. 2(a): Class 1: “more distance-sensitive” patient; 2(b): Class 2: “less distance-sensitive” patient.

**Table 3.**
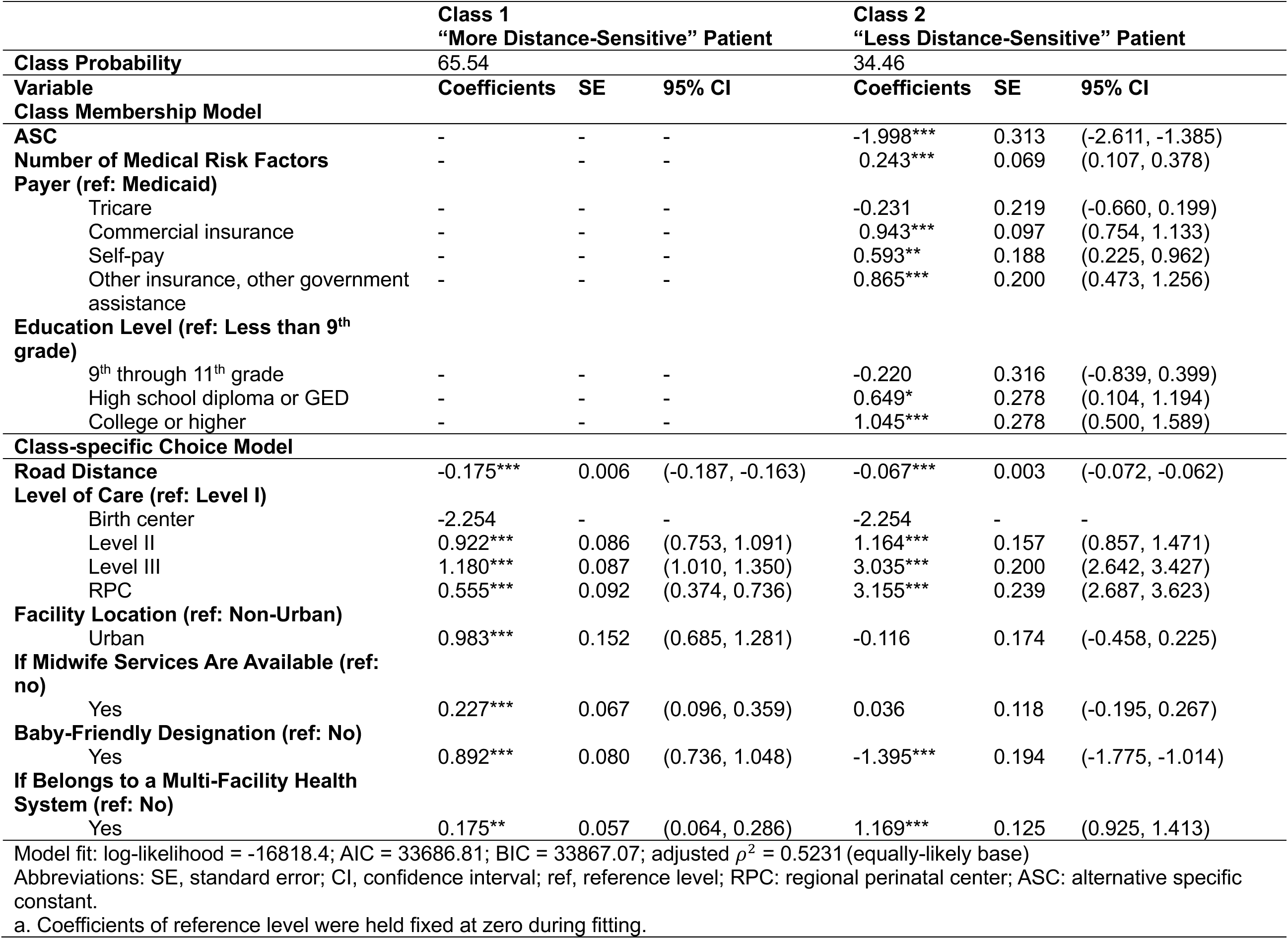
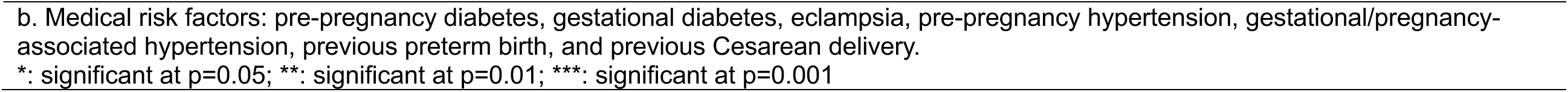
Estimated results for the final latent class model.

**Table 4.** Road distance equivalent of the fitted multinomial logit and latent class models.

| Variable | MNL<br>RDE (in miles) | LC class 1<br>RDE (in miles) | LC class 2<br>RDE (in miles) |
| --- | --- | --- | --- |
| **Level of Care (ref: Level I)** |  |  |  |
| Birth center | -20.39 | -12.89 | -33.74 |
| Level II | 6.79 | 5.27 | 17.43 |
| Level III | 13.21 | 6.74 | 45.43 |
| RPC | 9.09 | 3.17 | 47.24 |
| **Facility Location (ref: Non-Urban)** |  |  |  |
| Urban | 5.99 | 5.62 | 1.74 |
| **Midwife Service (ref: No)** |  |  |  |
| Yes | 1.54 | 1.30 | 0.54 |
| **Baby-Friendly Designation (ref: No)** |  |  |  |
| Yes | 2.14 | 5.10 | 20.88 |
| **Multi-facility Health System (ref: No)** |  |  |  |
| Yes | 3.76 | 1.00 | 17.50 |
| Abbreviations: RDE, road distance equivalent. |  |  |  |

The class membership model revealed patient characteristics associated with each latent class, as shown in **Table 3**. The model estimated that a randomly selected patient would fall into the “more distance-sensitive” class with a probability of 65.54% and “less distance-sensitive” with a probability of 34.46%. Patients with greater number of medical risk factors (estimate: 0.243, 95% CI: 9.107-0.378, p = 0.001), those with commercial insurance compared to those Medicaid-insured (estimate: 0.943, 95% CI: 0.754-1.133, p<0.001) and those who had a college degree or higher compared to those had less than 9^th^ grade (estimate: 1.045, 95% CI: 0.500-1.589, p<0.001) were more likely to be “less distance-sensitive”.

#### 3.3.2 Class profile

**Table 5** details class profiles for the two latent classes (Class 1: “more distance-sensitive” patients: 65.54%; Class 2: “less distance-sensitive” patients: 34.46%). We calculated expected values for patient-level characteristics. All variables included in the final LC class membership model showed distinct distribution across the two latent classes. Specifically, the expected number of medical risk factors was 0.354 for the “less-distance sensitive” group compared to 0.285 for the “more distance-sensitive” group. 54.4% of the “more distance-sensitive” group were Medicaid insured, while only 30.0% of the “less distance-sensitive” group were Medicaid insured. Compared to Class 2, Class 1 had higher rates of commercially insured patients (57.4% vs. 31.0%), patients with college or higher education (70.4% vs. 48.4%), and had lower rates of patients with inadequate or intermediate level of prenatal care utilization index (18.4% vs. 23.9%).

**Table 5.**
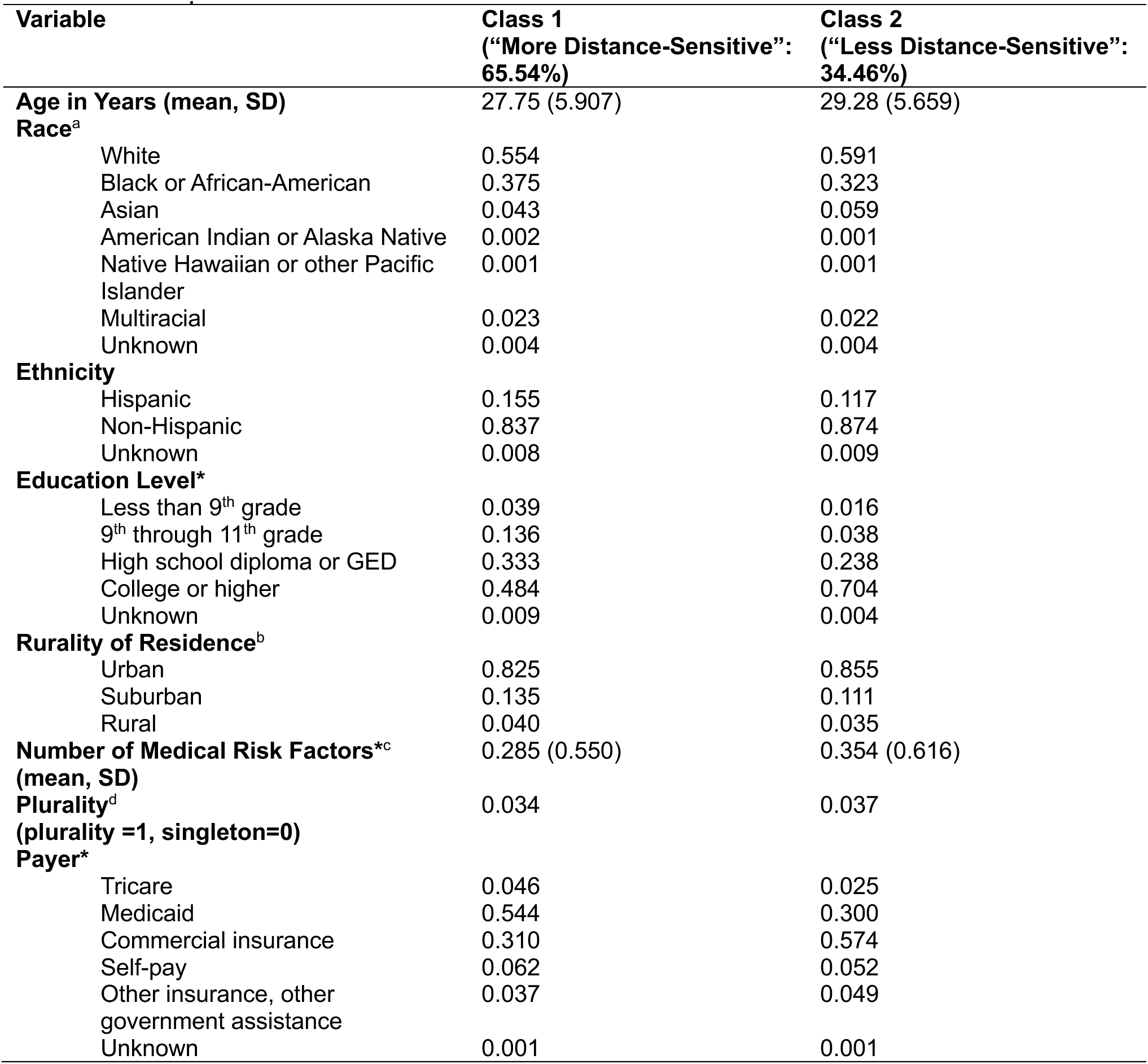
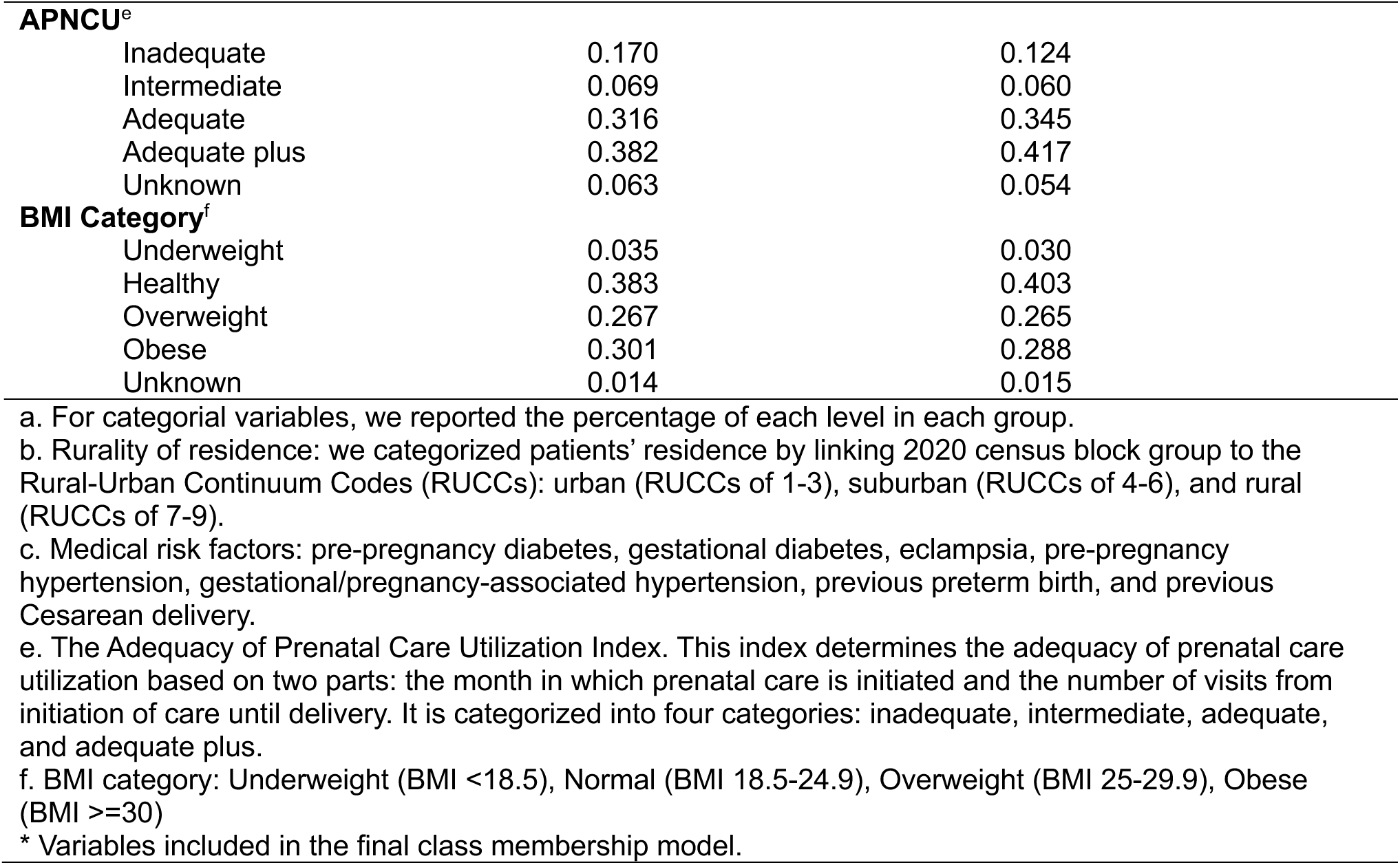
Class profiles of two latent classes.

| **Variable** | **Class 1**<br>**("More Distance-Sensitive":**<br>**65.54%)** | **Class 2**<br>**("Less Distance-Sensitive":**<br>**34.46%)** |
| --- | --- | --- |
| **Age in Years (mean, SD)** | 27.75 (5.907) | 29.28 (5.659) |
| **Race<sup>a</sup>** |  |  |
| White | 0.554 | 0.591 |
| Black or African-American | 0.375 | 0.323 |
| Asian | 0.043 | 0.059 |
| American Indian or Alaska Native | 0.002 | 0.001 |
| Native Hawaiian or other Pacific<br>Islander | 0.001 | 0.001 |
| Multiracial | 0.023 | 0.022 |
| Unknown | 0.004 | 0.004 |
| **Ethnicity** |  |  |
| Hispanic | 0.155 | 0.117 |
| Non-Hispanic | 0.837 | 0.874 |
| Unknown | 0.008 | 0.009 |
| **Education Level\*** |  |  |
| Less than 9<sup>th</sup> grade | 0.039 | 0.016 |
| 9<sup>th</sup> through 11<sup>th</sup> grade | 0.136 | 0.038 |
| High school diploma or GED | 0.333 | 0.238 |
| College or higher | 0.484 | 0.704 |
| Unknown | 0.009 | 0.004 |
| **Rurality of Residence<sup>b</sup>** |  |  |
| Urban | 0.825 | 0.855 |
| Suburban | 0.135 | 0.111 |
| Rural | 0.040 | 0.035 |
| **Number of Medical Risk Factors\*<sup>c</sup>**<br>**(mean, SD)** | 0.285 (0.550) | 0.354 (0.616) |
| **Plurality<sup>d</sup>**<br>**(plurality =1, singleton=0)** | 0.034 | 0.037 |
| **Payer\*** |  |  |
| Tricare | 0.046 | 0.025 |
| Medicaid | 0.544 | 0.300 |
| Commercial insurance | 0.310 | 0.574 |
| Self-pay | 0.062 | 0.052 |
| Other insurance, other<br>government assistance | 0.037 | 0.049 |
| Unknown | 0.001 | 0.001 |
| **APNCU<sup>e</sup>** |  |  |
| Inadequate | 0.170 | 0.124 |
| Intermediate | 0.069 | 0.060 |
| Adequate | 0.316 | 0.345 |
| Adequate plus | 0.382 | 0.417 |
| Unknown | 0.063 | 0.054 |
| **BMI Category<sup>f</sup>** |  |  |
| Underweight | 0.035 | 0.030 |
| Healthy | 0.383 | 0.403 |
| Overweight | 0.267 | 0.265 |
| Obese | 0.301 | 0.288 |
| Unknown | 0.014 | 0.015 |

a. For categorial variables, we reported the percentage of each level in each group.
b. Rurality of residence: we categorized patients' residence by linking 2020 census block group to the Rural-Urban Continuum Codes (RUCCs): urban (RUCCs of 1-3), suburban (RUCCs of 4-6), and rural (RUCCs of 7-9).
c. Medical risk factors: pre-pregnancy diabetes, gestational diabetes, eclampsia, pre-pregnancy hypertension, gestational/pregnancy-associated hypertension, previous preterm birth, and previous Cesarean delivery.
e. The Adequacy of Prenatal Care Utilization Index. This index determines the adequacy of prenatal care utilization based on two parts: the month in which prenatal care is initiated and the number of visits from initiation of care until delivery. It is categorized into four categories: inadequate, intermediate, adequate, and adequate plus.
f. BMI category: Underweight (BMI <18.5), Normal (BMI 18.5-24.9), Overweight (BMI 25-29.9), Obese (BMI ≥30)
\* Variables included in the final class membership model.

We also compared the expected values of sociodemographic variables for members of each class and found slight differences among the classes in terms of age, race, and ethnicity. Older, White, Asian, and non-Hispanic patients were more likely to be “less distance-sensitive”, while younger, Black or African-American, and Hispanic patients were more likely to be “more distance-sensitive”.

### 3.4 Out-of-sample Validation

#### 3.4.1 Choice prediction

We validated the models’ ability to predict individual choice of delivery facility (**Table 6**). LC model yielded a higher logarithmic score of -1.73, compared to a logarithmic score of -1.77 by MNL model and a logarithmic score of -35.54 by the distance-only model.

**Table 6.**
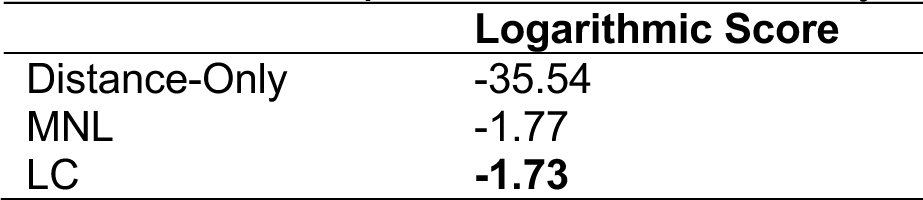
Choice prediction measured by logarithmic score.

#### 3.4.2 Forecast low-volume facilities

**Figure 3** shows true volume vs. volume predicted by the LC model in 2019 by volume decile. We also calculated and compared the LC model’s predictive performance with the distance-only model and the MNL model in forecasting low-volume facilities as shown in **Table 7**. The LC model consistently outperformed the distance-only and MNL models in terms of accuracy, sensitivity, and F1 score across all thresholds. At thresholds of 250, 300, and 400, the LC model achieved the highest F1 scores of 57.1, 66.7, and 81.3, respectively, compared to lower scores from the other models. At the threshold of 500, the LC model led in all metrics, with an accuracy of 93.5, sensitivity of 89.5, and F1 score of 87.2.

**Figure 3.**
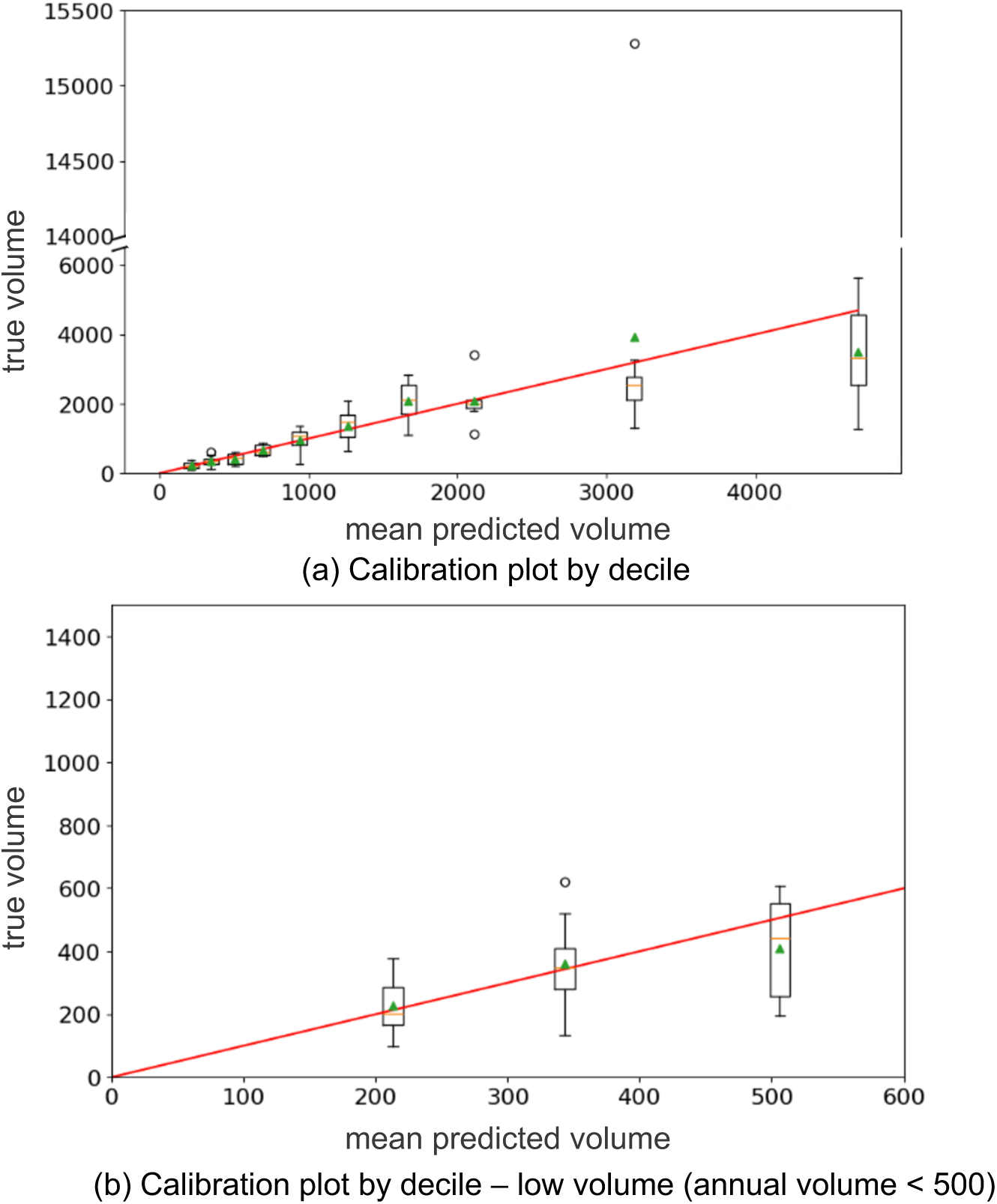
Calibration plots by decile comparing true annual volume vs. predicted volume from the latent class model on the 2019 data: (a) shows calibration plot by decile for all facilities; (b) shows calibration plot by decile for predicted low-volume facilities (decile mean < 500). Each bar is positioned at its mean value for the corresponding decile. Green triangles indicate decile mean; orange lines indicate decile median; the red line represents y=x line (perfect calibration).

**Table 7.**
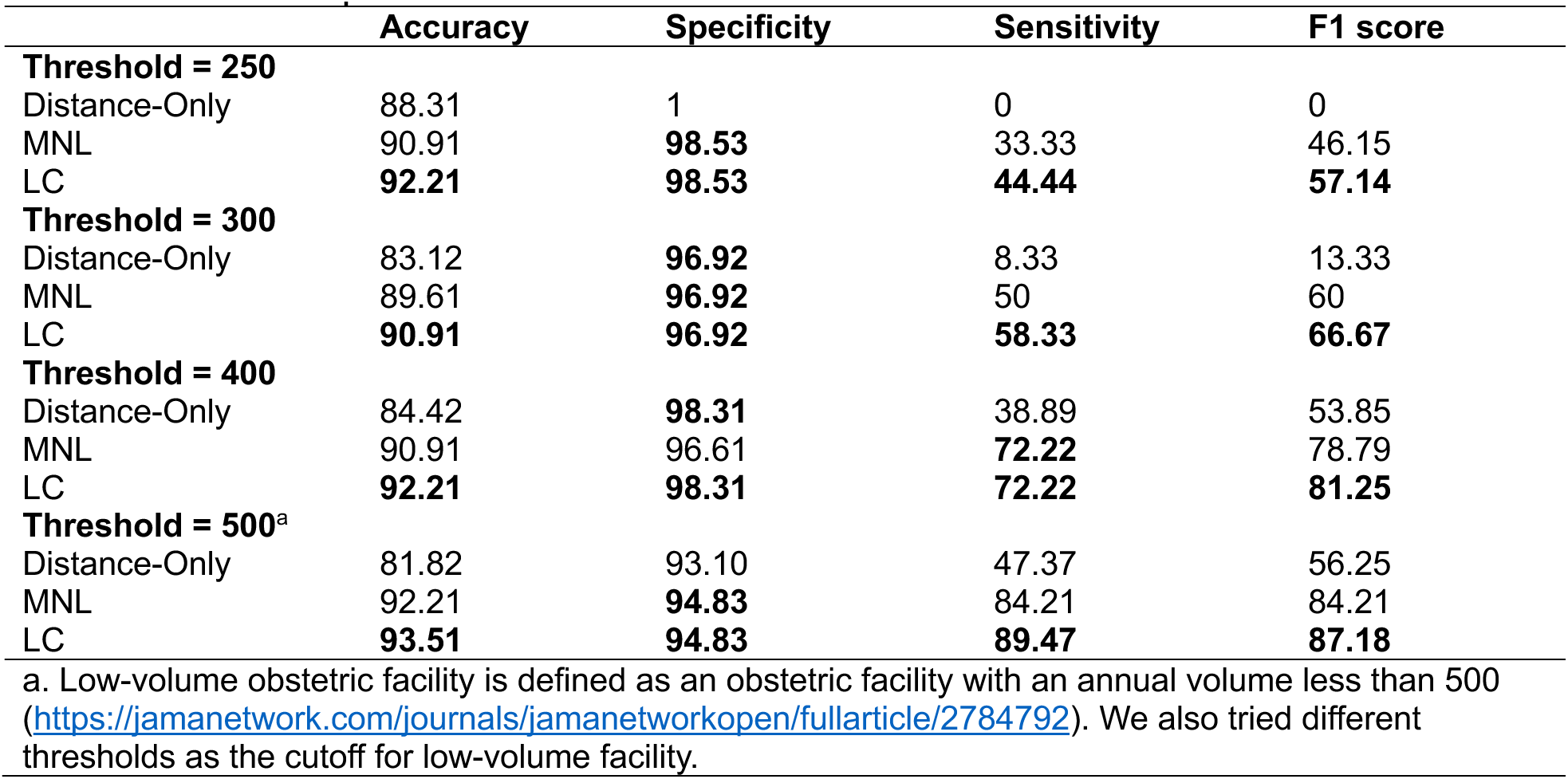
Predictive performance of low-volume facilities.

|  | Accuracy | Specificity | Sensitivity | F1 score |
| --- | --- | --- | --- | --- |
| **Threshold = 250** |  |  |  |  |
| Distance-Only | 88.31 | 1 | 0 | 0 |
| MNL | 90.91 | **98.53** | 33.33 | 46.15 |
| LC | **92.21** | **98.53** | **44.44** | **57.14** |
| **Threshold = 300** |  |  |  |  |
| Distance-Only | 83.12 | **96.92** | 8.33 | 13.33 |
| MNL | 89.61 | **96.92** | 50 | 60 |
| LC | **90.91** | **96.92** | **58.33** | **66.67** |
| **Threshold = 400** |  |  |  |  |
| Distance-Only | 84.42 | **98.31** | 38.89 | 53.85 |
| MNL | 90.91 | 96.61 | **72.22** | 78.79 |
| LC | **92.21** | **98.31** | **72.22** | **81.25** |
| **Threshold = 500<sup>a</sup>** |  |  |  |  |
| Distance-Only | 81.82 | 93.10 | 47.37 | 56.25 |
| MNL | 92.21 | **94.83** | 84.21 | 84.21 |
| LC | **93.51** | **94.83** | **89.47** | **87.18** |

a. Low-volume obstetric facility is defined as an obstetric facility with an annual volume less than 500 (https://jamanetwork.com/journals/jamanetworkopen/fullarticle/2784792). We also tried different thresholds as the cutoff for low-volume facility.

#### 3.4.3 Measure access to care

**Figures 4(a)-(d)** show the average travel distance to the chosen facility from their residence by census track, observed during the validation period (a) and predicted by our three models (b)-(d). The distance-only model predicted average travel distances with a Spearman correlation of 0.77 compared to observed values. Average travel distance predicted by both the MNL and the LC models aligned better with the observed pattern, as indicated by higher correlation coefficients of 0.83 and 0.84 between the observed average travel distance per census track and the average travel distance per census track (**Table 8**).

**Table 8.** Spearman correlation coefficients between observed and predicted metrics.

| Model | Average Travel Distance by<br>Census Track | Low Volume by Census<br>Track |
| --- | --- | --- |
| Distance-Only | 0.77 | 0.37 |
| MNL | 0.83 | 0.41 |
| LC | **0.84** | **0.42** |

**Figure 5(a)-(d)** show the percentage of patients who delivered at a low-volume facility with less than 500 annual birth volumes. All models identified similar regions where high percentage of patients received OB care at a facility predicted as a low-volume facility by each model. We observed that predicted pattern by the LC model again aligned the best with the observed pattern among all fitted models, indicated by the highest Spearman correlation coefficient of 0.42 between the observed percentage and the predicted percentage by the LC model (**Table 8**).

## 4. Discussion

In this article, we demonstrated a discrete choice modeling approach to estimate patients’ realized access to delivery facilities. Our findings suggested that distance, facility level of care, location of facilities, and affiliation with a multi-facility health system significantly affected patients’ selection of delivery location. Importantly, our study emphasized the importance of accounting for patient heterogeneity when evaluating access to care and designing policies to improve OB service delivery. Through a latent class analysis, we identified a small subgroup of “less distance-sensitive” patients (34.46%) who exhibited a stronger preference for facilities with higher levels of care relative to road distance to facilities compared to the majority of the “more distance-sensitive” group (65.54%). Our final LC model achieved an adjusted *ρ*\* of 0.52, which exceeded the conventional threshold of 0.4 for a good model fit [62]. Additionally, we evaluated the predictive performance of our models using out-of-sample data. Fitted models were applied to forecast patients’ facility choices, identify hospitals at risk of low delivery volumes, and evaluate maternal care access under two key measures: (1) average travel distance to a birthing facility and (2) the proportion of deliveries occurring in predicted low-volume hospitals. The LC model that incorporated both facility attributes and patient characteristics outperformed other models across every evaluation metric.

The smaller subgroup of “less distance-sensitive” patients who placed high preference for facilities with higher level and specialized care exhibited characteristics consistent with patterns reported in prior studies. Our findings align with evidence that patients with higher socioeconomic status were more likely to bypass nearby facilities in favor of facilities perceived to provide higher quality care [7, 17, 63]. Interestingly, while this subgroup showed higher utilization of facilities with advanced levels of care, they were less likely to deliver at “Baby-Friendly” designated hospitals. Although Baby-Friendly designation is intended to promote breastfeeding and maternal-infant bonding, hospitals with such designation were relatively rare in Georgia during the study period (only 5 out of 77 facilities from 2016 to 2019) and may not have been co-located with facilities offering higher levels of maternal and neonatal care. For patients with multiple medical risk factors and needing more advanced care, Baby-Friendly facilities may not have met their clinical needs.

Our study adds to the literature by providing insights into hospital choices and OB bypassing. We identified factors associated with choosing a facility farther away and quantified the tradeoffs patients make between facility attributes and travel distance. These findings have important policy implications. First, the strong influence of socioeconomic status on patients’ selection of birthing facilities emphasizes the need to design policy interventions that address patients’ social determinants of health. Simply expanding high-quality maternal care will not ensure equitable access if systemic barriers, such as transportation limitations, restrictive insurance networks, and health literacy gaps remain unaddressed. Additionally, our results confirm the importance of risk-appropriate care where patients should seek care at a facility that is suited to handle their risk. Matching patients to facilities with appropriate levels of care is critical to reducing adverse maternal outcomes, as high-risk deliveries at lower-level facilities are associated with poorer outcomes. However, availability of higher quality services cannot impact maternal outcomes if women do not choose to give birth in high-quality facilities. Policymakers should focus on mechanisms that facilitate appropriate matching between patient risk and facility capability, such as regionalized care systems, referral pathways, and patient navigation programs

Our proposed measure of access to care, based on the percentage of patients delivering at low-volume facilities, offers a different perspective on access to maternal care amid OB unit closures. While measures like travel distance to delivery facilities (such as Maternity Care Deserts or distance to CCO services) and others (such as regions with high maternal transport rate) identified similar at-risk regions in central east Georgia, these areas have a low percentage of patients delivering at low-volume facilities [15, 16, 64–66]. This highlights the trade-off between keeping more OB units open to reduce geographic barriers and the potential for lower OB volume due to spreading out service providers. Policymakers must weigh the benefits of proximity against the risks associated with low-volume care, particularly in rural regions. If distance were the main factor in patients’ choice of delivery facilities, we would expect the number of low-volume facilities predicted by a distance-only model to match the actual number. However, the predicted number was lower when distance was the only factor considered. This confirms the risk of patients bypassing nearby facilities for more appealing ones, potentially leaving local facilities with low volumes.

Our discrete choice modeling framework provides a practical, data-driven tool for identifying facilities at risk of underutilization and informing policies to better distribute patient volume across the region. For example, a Level I facility in Northeast Georgia, which was only correctly identified as low-volume by both our MNL and LC models by taking the realized access perspective, later confirmed to have closed its OB unit in 2025. In contrast, the distance-only model which reflects potential rather than actual access, failed to identify this facility as underutilized. Our discrete choice models are also highly adaptable and can be incorporated in other advanced models for future policy analysis. One such use case is demonstrated in Meredith and Steimle 2025, where a choice model is incorporated in an optimization framework to identify regions that were vulnerable to closures [67].

There are opportunities to further improve the modeling approaches described in this study. First, although LC model demonstrates superior predictive performance compared to the distance-only and MNL models, our model underestimated failed to capture a significant proportion of delivery volume at the facility with the highest annual volume. Additionally, rurality of patients’ residence was not chosen in our final LC model, counter-intuitive to the belief that rural and urban patients would have different preferences for healthcare facilities. We initially included this variable into LC model, but this variable turned out to be insignificant or the model that included this variable had worse fit and predictive performance. We suspect that the difference of facility choices between urban and rural residents could be explained by other factors, such as distance to the facility, patients’ socioeconomic characteristics, and payers.

Our study has several limitations. First, Georgia birth records do not include deliveries of out-of-state residents in Georgia facilities. This omission could underestimate delivery volumes at facilities near state borders, although the proportion of out-of-state residents delivering in Georgia facilities is expected to be small. Because our modeling population included only Georgia residents, the absence of out-of-state patients from the dataset should not bias our estimates of realized access to birthing facilities, as those patients were not part of the modeling population. Future research can consider linking birth records with data from the American Hospital Association to obtain accurate annual OB volume by both in-state and out-of-state residents. Second, we used driving distance as a measure of proximity from patients’ residences to OB facilities. This approach could underestimate barriers to OB access for those with low socioeconomic status, as they might lack access to a car or reliable transportation. Additionally, our analysis is limited to deliveries that occurred at OB facilities within 120 miles from the birthing person’s residence. Future research should examine non-facility births, deliveries occurring at non-OB facilities, and births taking place far from the patient’s residence, as these cases may reveal additional barriers to accessing facility-based OB care. In particular, studying patients who travel exceptionally long distances for specialized care could provide insights into access challenges faced by individuals with complex medical needs or limited local options for high-risk OB care. Lastly, our analysis relied on RP to estimate patients’ choices. This method does not allow us to differentiate between active choices, where patients deliberately select a facility, and passive choices, where external constraints such as availability or referral patterns dictate the decision.

In conclusion, patients placed heterogeneous weights on OB facility attributes. Patients who have more severe medical conditions, have commercial insurance compared to Medicaid, and are highly educated are more likely to be “less distance-sensitive” and select facilities with higher levels of care more often. Incorporating this heterogeneity into predictive models improves the identification of communities at risk, specifically, areas where a high proportion of patients must travel long distances not by choice or deliver at low-volume facilities. Efforts to improve maternal care access should go beyond addressing geographic barriers alone. Effective policies aimed at enhancing access should also consider broader social determinants, including differences in insurance coverage and educational attainment, which affect patients’ ability and willingness to reach higher-quality care. Lastly, our study emphasizes the importance of a realized access perspective when measuring access to care. Our model can serve as a practical tool for identifying underutilized facilities and informing future policies that better distribute patient volume across the region.

## Statements and Declarations

### Data Availability

We are not able to share Georgia birth records data due to our data use agreement.

### Ethics Approval

This study was conducted in accordance with the ethical principles outlined in the Declaration of Helsinki (https://www.wma.net/policies-post/wma-declaration-of-helsinki/). IRB approval was obtained from the Georgia Institute of Technology (Protocol H23091), and the need for consent was waived by the IRB.

### Authors’ Contributions

Jingyu Li: conceptualization, data curation, methodology, formal analysis, validation, writing – original draft, visualization.

Margaret Carrel: validation, writing – review and editing.

Stephanie M. Radke: validation, writing – review and editing.

Lauren N. Steimle: data acquisition, conceptualization, methodology, writing – original draft, project administration, funding acquisition, supervision.

## Acknowledgements

We want to thank Dr. Stephane Hess, the co-author of the Apollo package, for his help with the initial software setup and troubleshooting. We also want to thank the Georgia Department of Public Health for providing birth records data.

