## Supplemental Information for "Estimating Realized Access to Obstetric Care in Georgia: A Discrete Choice Modeling Analysis"

#### Appendix A. Data Processing

Table A1. Description of variables

| Variable | Description | Type | Values |
|---|---|---|---|
| Age in years | The mother's age in years at the time of delivery. | Numeric | range 10-55 years |
| Race | White = a person having origins in any of the original peoples of Europe, the Middle East or North Africa; Black or African-American = A person having origins in any of the black racial groups of Africa; Asian=A person having origins in any of the original peoples of the Far East, Southeast Asia, or the Indian subcontinent including for example, Cambodia, China, India, Japan, Korea, Malaysia, Pakistan, the Philippine Islands, Thailand and Vietnam; American Indian/Alaska Native=A person having origins in any of the original peoples of North and South America (including Central American), and who maintains tribal affiliation or community attachment; Native Hawaiian or Other Pacific Islander=A person having origins in any of the original peoples of Hawaii, Guam, Samoa, or other Pacific Islands. Multiracial = 2 or more of these races (OMB-15, 1997). | Categorical | White, Black or African-American, Asian, American Indian/Alaska Native, Native Hawaiian or Other Pacific Islander, Multiracial |
| Ethnicity | Ethnicity, currently limited to asking whether the person is “Hispanic or Latino” (A person of Mexican, Puerto Rican, Cuban, South or Central American, or other Spanish culture or origin, regardless of race) (OMB-15, 1997). | Categorical | Hispanic, Non-Hispanic |
| Rurality of residence | Patients’ residence by linking 2020 census block group to the Rural-Urban Continuum Codes (RUCCs): urban (RUCCs of 1-3), suburban (RUCCs of 4-6), and rural (RUCCs of 7-9). | Categorical | Urban, Suburban, Rural |

| | | | |
|---|---|---|---|
| Education level | The last grade of formal education completed. | Categorical | Less than 9th Grade, 9th through 11th Grade, High School Diploma or GED (12), Some College or Higher |
| Payer | Principal source of payment | Categorical | Commercial Insurance, Medicaid, Medicaid Applicants, or Medicaid Managed Care, Other Insurance, Government Assistance, or Other Non-specified Managed Care, Self-Pay, TRICARE |
| APNCU index | the Adequacy of Prenatal Care Utilization Index | Categorical | Inadequate, Intermediate, Adequate, Adequate plus |
| Number of medical risk factors | Number of medical risk factors patient has among: pre-pregnancy diabetes, gestational diabetes, eclampsia, pre-pregnancy hypertension, gestational/pregnancy-associated hypertension, previous preterm birth, and previous Cesarean delivery. | Numeric | range 0-7 |
| Plurality | Number of fetuses for this pregnancy. | Numeric | range 1-8 |

Table A2. Variance inflation factors for candidate facility-level attributes

| **Variable** | $GVIF$ | $DF$ | $GVIF^{\frac{1}{2DF}}$ [a] |
|---|---|---|---|
| Level of Care (ref: Level I) | 1.68 | 4 | 1.07 |
| Facility Location (ref: non-urban) | 1.80 | 1 | 1.34 |
| If midwife services are available (ref: no) | 1.37 | 1 | 1.17 |
| If belongs to a multi-facility health system (ref: no) | 1.38 | 1 | 1.17 |

Abbreviations: GVIF, generalized variance inflation factor; DF: degree of freedom.

a. We reported $GVIF^{\frac{1}{2DF}}$ to make $GVIF$ values comparable across variables with different degrees of freedom.

Table A3. MNL estimates using 10000 sampled data

| Variable | Coefficients | SE | 95% CI |
|---|---|---|---|
| **Road Distance** | -0.109*** | 0.001 | (-0.111, -0.107) |
| **Level of Care (ref: Level I)**[a] | | | |
| Birth center | -2.346*** | 0.241 | (-2.819, -1.874) |
| Level II | 0.758*** | 0.051 | (0.658, 0.858) |
| Level III | 1.482*** | 0.051 | (1.383, 1.581) |
| RPC | 1.101*** | 0.055 | (0.994, 1.208) |
| **Facility Location (ref: Non-Urban)** | | | |
| Urban | 0.771*** | 0.068 | (0.637, 0.905) |

Model fit: log-likelihood = -17462.57; AIC = 34937; BIC = 34980; adjusted $\rho^2$ = 0.5054 (equally-likely base)

Abbreviations: SE, standard error; CI, confidence interval; ref, reference level; RPC: regional perinatal center.

a. Coefficients of reference level are held fixed at zero during fitting.

*: significant at p=0.05; **: significant at p=0.01; ***: significant at p=0.001

Table A4. MNL estimates using 2016-2018 data

| Variable | Coefficients | SE | 95% CI |
|---|---|---|---|
| **Road Distance** | -0.111*** | 0.0002 | (-0.111, -0.1105) |
| **Level of Care (ref: Level I)**[a] | | | |
| Birth center | -2.161*** | 0.036 | (-2.232, -2.090) |
| Level II | 0.744*** | 0.009 | (0.727, 0.760) |
| Level III | 1.473*** | 0.008 | (1.456, 1.489) |
| RPC | 1.087*** | 0.009 | (1.070, 1.105) |
| **Facility Location (ref: Non-Urban)** | | | |
| Urban | 0.797*** | 0.011 | (0.774, 0.819) |

Model fit: log-likelihood = -644152.3; AIC = 1288317; BIC = 1288381; adjusted $\rho^2$ = 0.51 (equally-likely base)

Abbreviations: SE, standard error; CI, confidence interval; ref, reference level; RPC: regional perinatal center.

a. Coefficients of reference level are held fixed at zero during fitting.

*: significant at p=0.05; **: significant at p=0.01; ***: significant at p=0.001

### Appendix B. Additional Modeling Results

Table B1. Model fit of 3-class LC model

| Log-Likelihood | AIC | BIC | adjusted $\rho^2$ |
|---|---|---|---|
| -16776.9 | 33879.38 | 34146.17 | 0.52 |

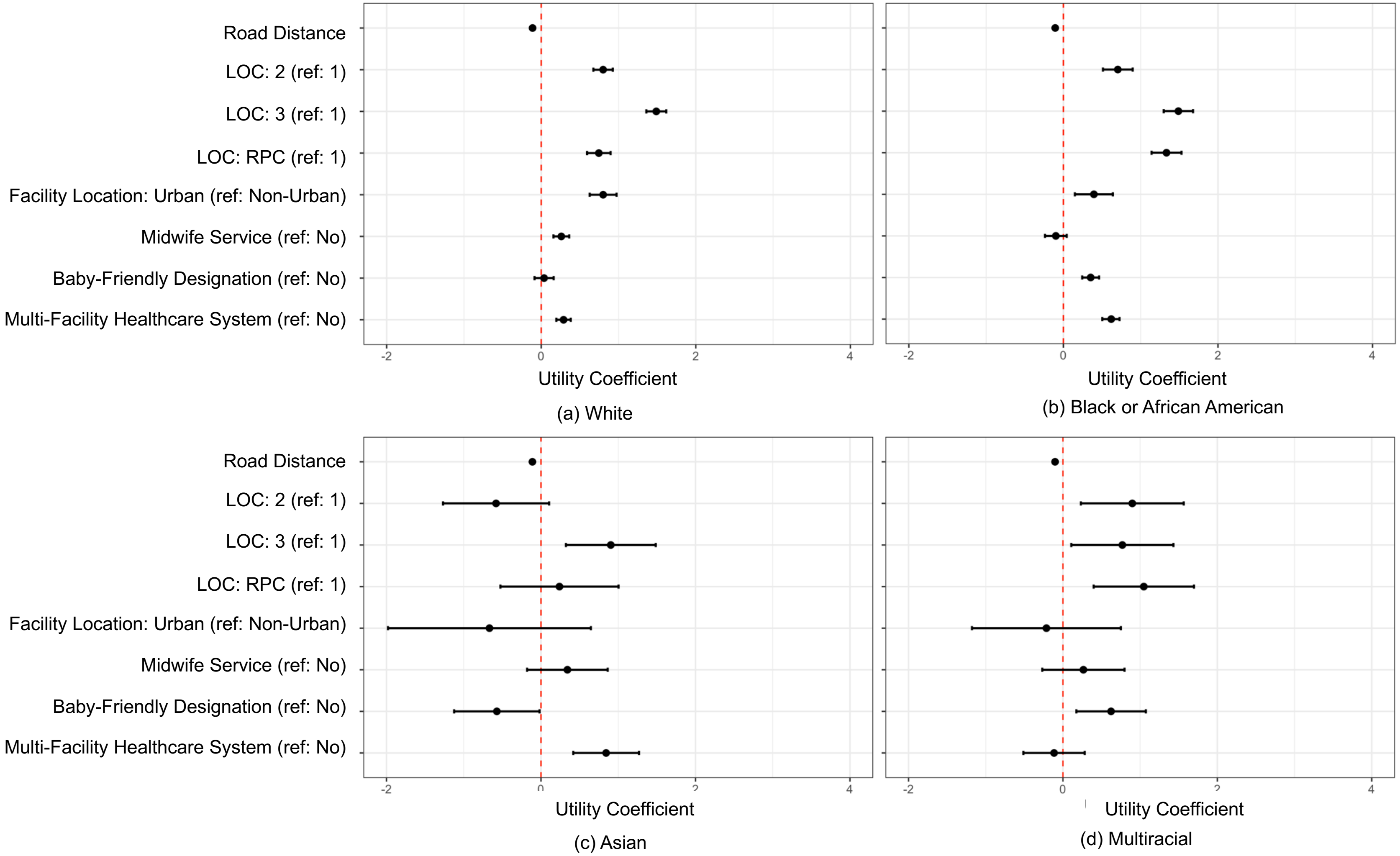


Figure B1. MNL estimates for subgroup analysis of race. A1(a) White; A1(b) Black or African American; A1(c) Asian; A1(d) Multiracial.

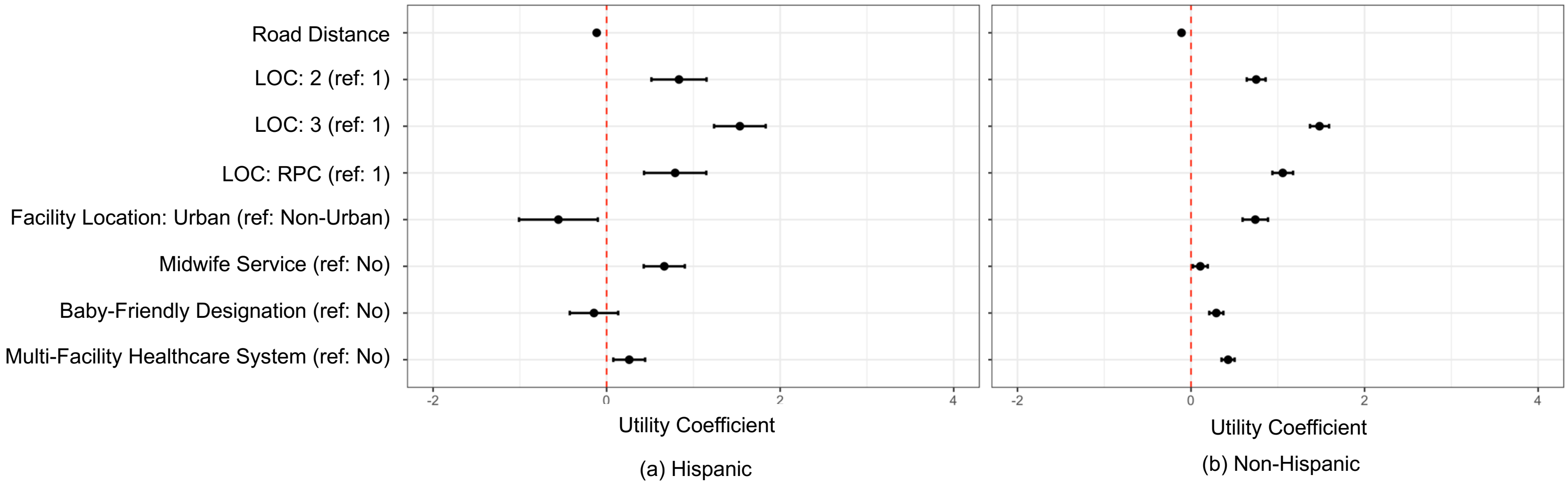


Figure B2. MNL estimates for subgroup analysis of ethnicity. A2(a) Hispanic; A2(b) Non-Hispanic.

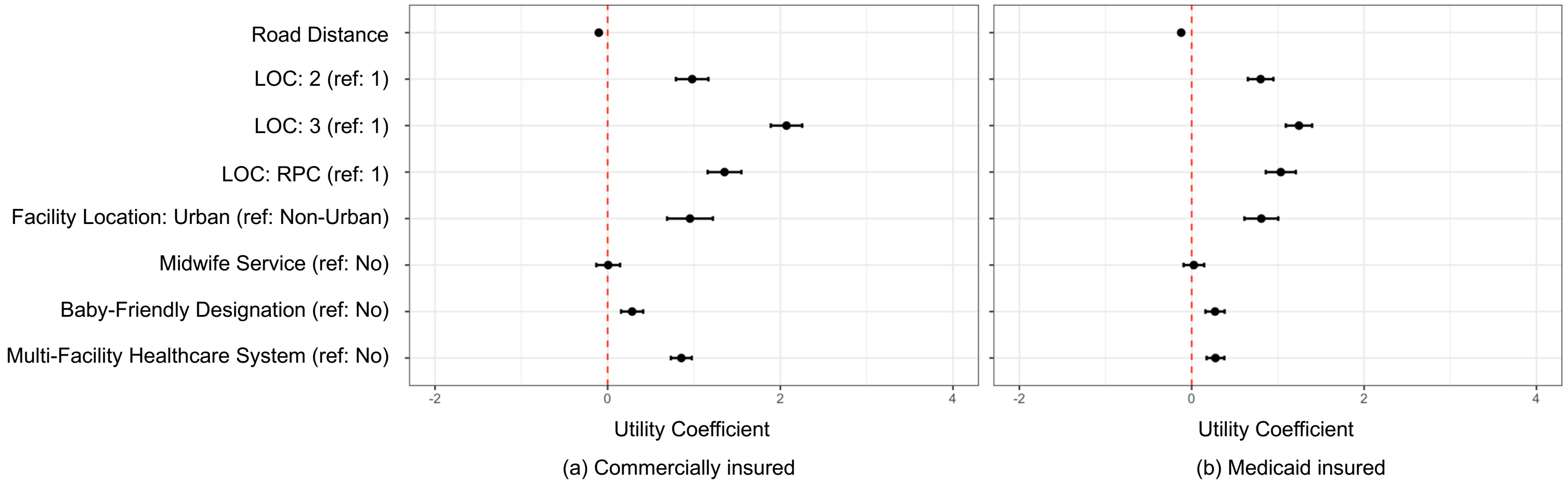


Figure B3. MNL estimates for subgroup analysis of payer status. A3(a) Commercially insured; A3(b) Medicaid insured.

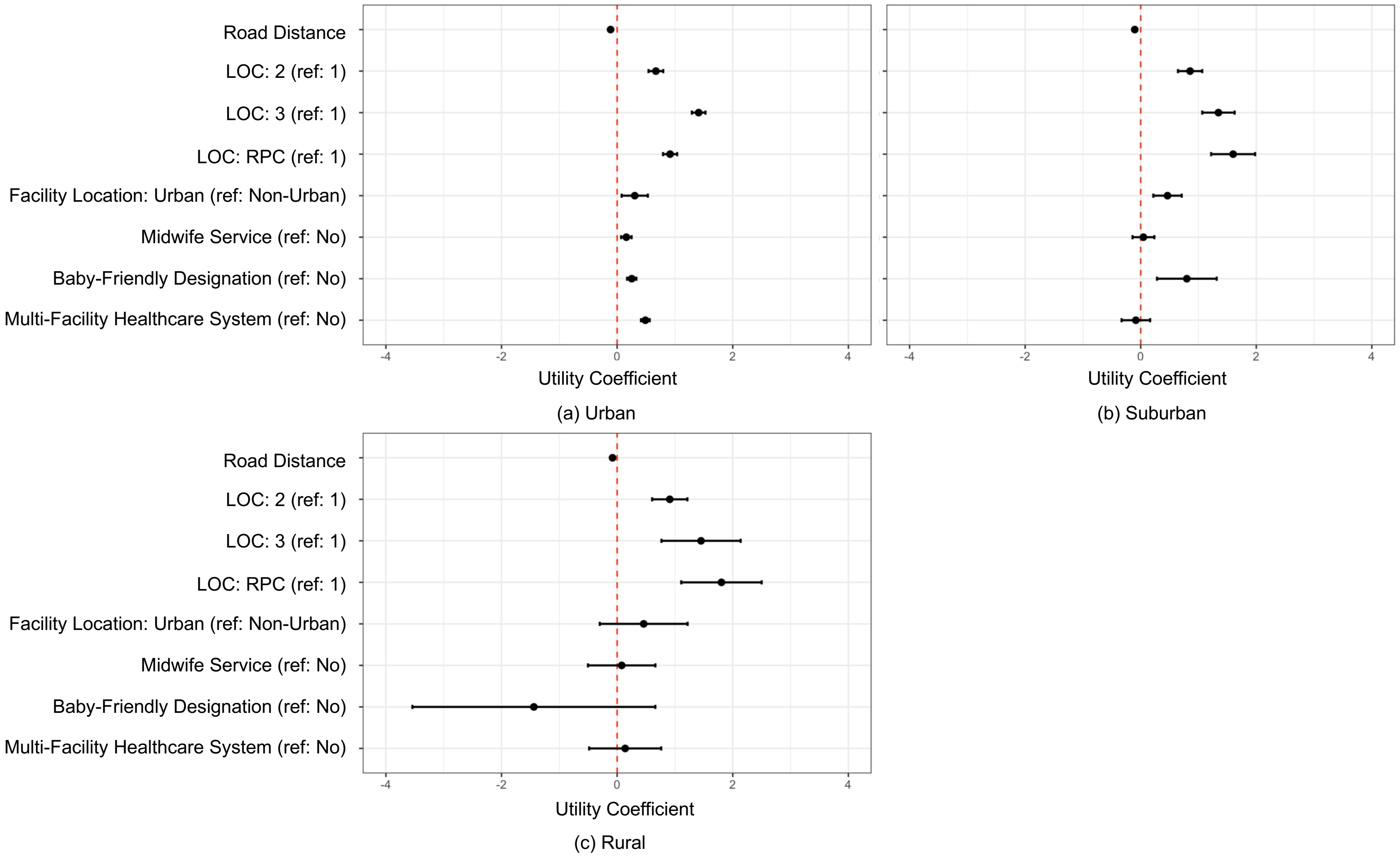


Figure B4. MNL estimates for subgroup analysis of rurality of residence. A4(a) Urban;A4(b) Suburban; A4(c) Rural.

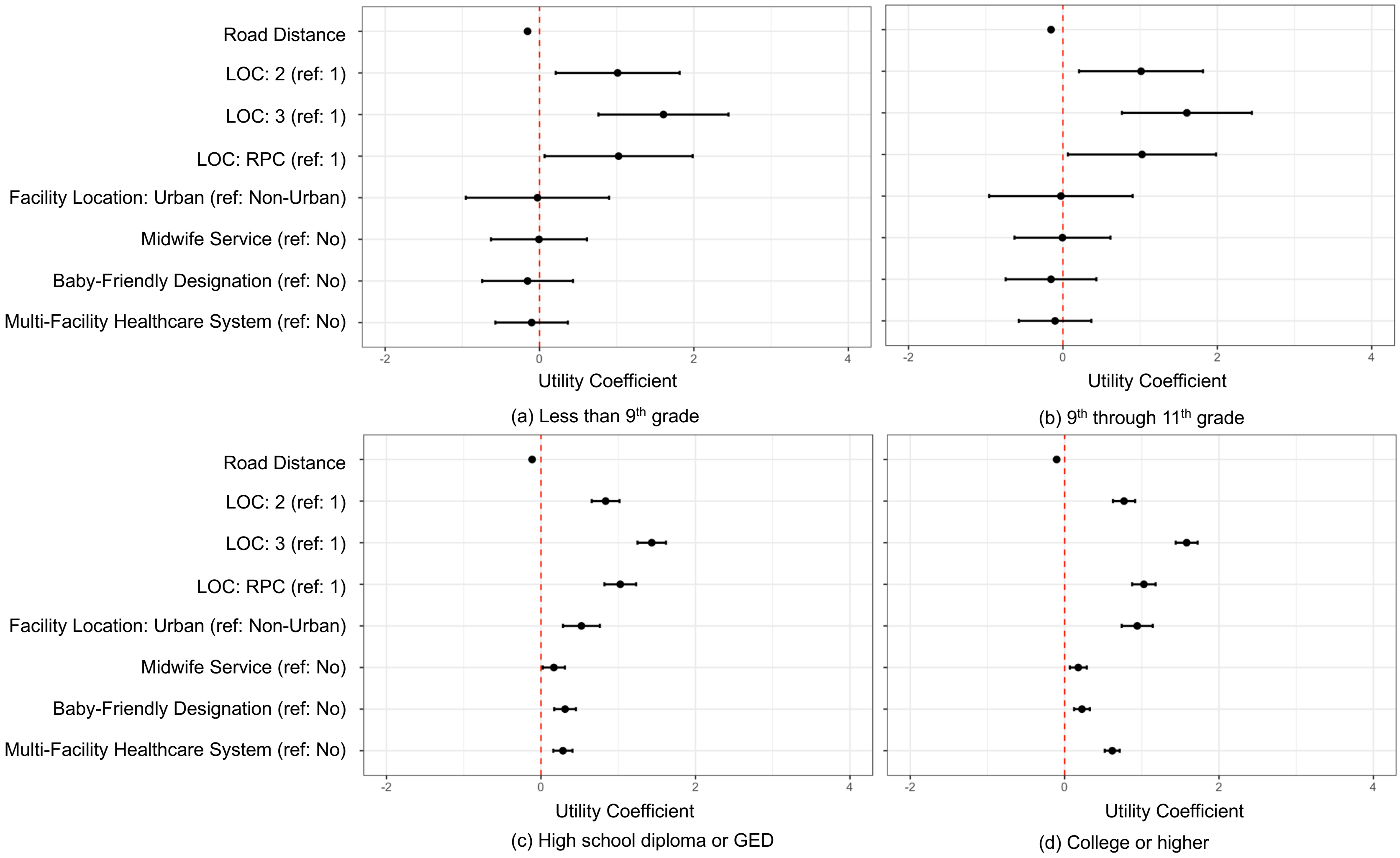


Figure B5. MNL estimates for subgroup analysis of education level. A5(a) Less than 9th grade; A5(b) 9th through 11th grade; A5(c) High school diploma or GED; A5(4) College or higher.
